# Topographical overlapping of the Aβ and Tau pathologies in the Default mode networks predicts Alzheimer’s Disease with higher specificity

**DOI:** 10.1101/2021.03.09.21253175

**Authors:** Seyed Hani Hojjati, Farnia Feiz, Sindy Ozoria, Qolamreza R. Razlighi, for the Alzheimer’s Disease Neuroimaging Initiative

**Author notes:** Data used in preparation of this article were obtained from the Alzheimer’s Disease Neuroimaging Initiative (ADNI) database (adni.loni.usc.edu). As such, the investigators within the ADNI contributed to the design and implementation of ADNI and/or provided data but did not participate in analysis or writing of this report. A complete listing of ADNI investigators can be found at: http://adni.loni.usc.edu/wp-content/uploads/how_to_apply/ADNI_Acknowledgement_List.pdf.

## Abstract

While Amyloid-plaques and Tau-tangles are the well-recognized pathologies of Alzheimer’s disease (AD), they are more often observed in healthy individuals than in AD patients. This discrepancy makes it extremely challenging to utilize these two proteinopathies as reliable biomarkers for the early detection as well as later diagnosis of AD. Using the recent advancements in the imaging technology, our newly developed quantification methods, and publicly available neuroimaging data from 303 individuals, we hypothesize and provide preliminary evidence that topographically overlapping Aβ and Tau within the DMN play more critical roles in the underlying pathophysiology of AD than the Tau and/or Aβ pathologies. We first showed that the probability of observing overlapping Aβ and Tau is significantly higher within the DMN than outside DMN. Then we showed evidence that using Aβ and Tau overlap can increase the reliability of the prediction of healthy individuals converting to MCI and a lesser degree converting from MCI to AD. These findings shed some light on the complex pathophysiology of AD and suggest that overlapping Aβ and Tau pathologies within DMN might be a more reliable biomarker of AD for early detection and later diagnosis of the disease.

## Introduction

Alzheimer’s disease (AD) is the most common cause of dementia in older adults and has high morbidity and mortality [1]. An estimated 5 million people over the age of 65 in the US live with AD and are projected to rise to 13.8 million in the US and more than 130 million worldwide by 2050 [2]. AD is a slowly evolving disorder that usually reveals the dysfunction of the memory system and leads to the irreversible impairment of other cognitive functions [3]. The clinical/behavioral symptoms include episodic memory impairment (the most common initial symptom), and later symptoms of impaired cognitive function, and neuropsychiatric symptoms [4, 5]. However, disease progression can vary between 5 to 20 years to reach an endpoint where early hints of memory deficits can initially appear. In fact, many patients reach their end of life without developing fully characterized AD [4]. This makes the diagnosis of AD extremely challenging and leads to an ambiguous and constantly changing diagnostic criterion for identifying AD. Therefore, current diagnostic criteria for AD rely significantly on the imaging biomarkers of the AD pathologies [6-9]. Despite such high prevalence, the exact pathogenesis of AD is still under investigation. The aggregation of two misfolded proteins - intracellular neurofibrillary Tau tangles and extracellular amyloid-β (Aβ) plaques - are the best-documented pathologies for AD [10, 11]. However, there is a long asymptomatic period (up to two decades) between the onset of pathologic changes in the brain and the earliest development of clinical symptoms of AD [12]. Thus, the biological processes underlying AD occurs while the patient’s cognitive scores are still normal and are present for decades before the emergence of any symptoms affecting cognitive functions. Discrepancies between normal aging pathologies and age-related neurodegenerative diseases like AD have been questioned for decades and still pose a major challenge in the diagnosis of AD as well as the early detection of it. Therefore, not only is the progression of AD exceedingly difficult to predict, but its early detection using well-recognized imaging biomarkers is also extremely challenging.

While the exact pathogenesis of AD remains unclear, the aggregation of two proteins appears to be common in all forms of AD [13]. Small aggregates of extracellular Aβ peptide plaques in the form of oligomers and the deposition of the hyperphosphorylated form of Tau protein (p-Tau) in neural cytoplasm that forms a major component of neurofibrillary tangles, appears to be two major proteinopathies involved in the pathogenesis of AD [14]. However, spatiotemporal discrepancies between these two proteinopathies are a key unresolved challenge in understanding the pathogenesis of AD. Spatially, Aβ is shown to be ubiquitously accumulated throughout the medial parietal, and prefrontal cortex where negligible Tau pathology was found. Inversely, aggregation of Tau pathology seems to follow a more region-specific pathway starting from the medial temporal lobe (MTL) and spreading to the inferior temporal gyrus where little or no Aβ accumulation was reported [15-18]. The temporal discrepancies are even more puzzling. While the *Amyloid cascade hypothesis* of AD places accumulation of Aβ at the start of the AD pathophysiology about 15 years earlier than the onset of clinical symptoms at age 65 and older, there is compelling evidence for the aggregation of Tau pathology in the locus coeruleus and transentorhinal cortex in about 20% of a healthy population in their early adult lifespan (20 to 40 years). In addition, while about 25% of the older population will never show any trace of Aβ in their brain (even at age 90 and higher), almost everyone at that age will have Tau aggregation. Despite these spatiotemporal discrepancies, it seems that throughout normal aging, early accumulation of Tau and Aβ are shown to be initiated independently in different regions of the brain and at different time points during the adult lifespan [13]; however, there is unequivocal evidence that their accumulation starts interacting with each other later in adulthood but even more strongly during the initial course of the AD and mild cognitive impairment (MCI) [19, 20]. Currently, the time and location of this interaction is an unknown inquiry. Our main goal in this study is to introduce DMN as the location of this interaction and hypothesis that overlapping Aβ and Tau pathology within DMN can be considered as a more reliable imaging biomarker for early detection of AD.

Numerous structural and functional measurements of the Default mode network (DMN) (functional connectivity (FC), negative BOLD response (NBR), baseline hyper/hypometabolism, and brain atrophy) have shown it to be implicated in normal aging, the pre-clinical stage of AD, mild cognitive impairment and AD [12, 21, 22]. By measuring the DMN’s FC and task-evoked NBR simultaneously, we have previously reported that the DMN has two dissociable functional levels that are topographically overlapping but have distinct roles in the functional architecture of this large-scale brain network [23]. There is evidence that Aβ pathology reduces the FC of the DMN. Separately, Tau pathology seems to disrupt NBR in the DMN but has negligible effects on the FC of the same regions. Naturally, when both Aβ and Tau spatially coincide within the DMN, *double-insult*, they disrupt the normal operations of both functional levels of the DMN causing a complete breakdown of the network, which potentially can initiate a cascade of events that results in AD. In contrast, a solo and/or spatially non-overlapping accumulation of Aβ and Tau has milder consequences on the DMN functionality. Our hypothesis, if true, will provide the underlying reason for the weak relationship between the Aβ deposition and cognition, whereas its deposition level is shown to be strongly related to the FC of the DMN.

In this study, we used our recently developed and validated quantification method and partial volume correction technique for reconstructing Aβ and Tau PET scans found in the Alzheimer’s disease neuroimaging initiative (ADNI) database to provide preliminary evidence for our introduced *double-insult* hypothesis. By using our quantifying techniques, we can accurately measure the extent of Aβ and Tau accumulation on the surface of the cerebral cortex of cognitively normal, MCI, and AD patients with an unprecedented spatial resolution (almost on a millimeter-by-millimeter scale). Achieving such spatial resolution is essential in accurately measuring the spatial overlap of the regions with both Aβ and Tau accumulation. We first show that in AD patients the probability of observing spatially coinciding Aβ and Tau pathologies within the DMN is much greater than the rest of the brain. Then, we show the superior reliability of overlapping pathologies within DMN for predicting the conversion from HC to MCI and less degree from MCI to AD. These pieces of evidence suggest that the overlapping Tau and Aβ pathology in the DMN regions might be a more reliable biomarker for early detection of AD as well as monitory the progress of the disease.

## Methods

### Participants

Data used in the preparation of this article were obtained from the Alzheimer’s Disease Neuroimaging Initiative (ADNI) database (adni.loni.usc.edu). The ADNI was launched in 2003 as a public-private partnership, led by Principal Investigator Michael W. Weiner, MD. The primary goal of ADNI has been to test whether serial magnetic resonance imaging (MRI), positron emission tomography (PET), other biological markers, and clinical and neuropsychological assessment can be combined to measure the progression of mild cognitive impairment (MCI) and early Alzheimer’s disease (AD). To be able to compute the overlapping accumulation of Tau and Aβ, we need the subjects to have both Amyloid and Tau scans as well as a high-resolution structural T1 weighted scan being acquired within one year. We have identified 159 healthy control (12 of them are the subjects who progress to MCI, healthy control converter), 127 MCI (14 of them are the subjects who progress to AD, MCI converter), and 17 AD participants that fit our requirements. 190 number of identified participants had readily scanned with resting-state fMRI. All participants gave their written consent to participate in the ADNI repository and all imaging protocols were approved by the local institutional review board of the site where the data have been collected. The MCI converter patients were converted to AD between 6 and 36 months. AD patients had a mini-mental state examination (MMSE) score of 20–26, a clinical dementia rating (CDR) of 0.5 or 1.0, and met the national institute of neurological and communicative disorders and stroke and the AD and related disorders association (NINCDS/ADRDA) criteria for probable AD. MCI patients had MMSE scores between 24 and 30, a memory complaint, objective memory loss measured by education adjusted scores on Wechsler memory scale logical memory II, a CDR of 0.5, absence of significant levels of impairment in other cognitive domains, essentially preserved activities of daily living, and an absence of dementia. The normal subjects were non-depressed, non-MCI, and non-demented, and had an MMSE score of 24–30 and a CDR close to zero.

### MRI acquisition parameters

Both T1-weighted structural scan and resting-state fMRI scans were obtained using ADNI standardized pulse sequence (http://adni.loni.usc.edu/methods/documents/mri-protocols/) in more than 55 ADNI sites using 3 Tesla calibrated and quality-controlled scanners. 6 minutes and 20-second MPRAGE scan with TR/TE= 2300/2.96 ms, TI=900 ms, matrix size=240×256, field of view=24×25.6 cm, and 170-208 sagittal slices with 1-1.2 mm thickness have been acquired. A 10-minutes of resting-state fMRI data has been collected using TR/TE = 3000/30 ms, flip angle=90-degree, matrix size=88×88, field of view= 22×22 cm, and 65 axial slices with 2.5-3.4mm thickness where the subject instructed to lay down with eyes open, stay as still as possible, and not to fall sleep. Scan quality was evaluated by the ADNI MRI quality control center at the Mayo Clinic to exclude “failed” scans because of motion, technical problems, or significant clinical abnormalities.

### PET imaging acquisition

According to standardized protocols, all ADNI site’s PET scanner should have an up-to-date calibration and normalization on the date of each imaging session. Florbetapir (AV-45) is the tracer used for imaging Amyloid-b plaques in the ADNI dataset. A 370 MBq (10 mCi +/- 10%) bolus injection of AV-45 will be administered through an intravenous catheter (saline should not be added to the dose prior to administration). Scanning starts by acquiring a low-dose computed tomography (CT) image for PET attenuation correction. Then, a dynamic 3D continuous brain PET imaging for 20-minutes (four 5-minute frames) will begin 50 minutes post-injection. The images will be reconstructed immediately after the 20 minutes scan, and if a motion artifact is detected, another 20 minutes continuous scan will be acquired.

The 18F-AV1451 tracer was used for imaging the neurofibrillary Tau tangles in ADNI. PET imaging was performed at each ADNI site according to standardized protocols. A 370 MBq (10.0 mCi ± 10%) of AV1451 tracer was administered through an intravenous catheter. Dynamic 3D data were continuously acquired as six 5-minutes frames from 80 to 100 min post-tracer injection. A low-dose CT scan preceded these acquisitions for PET attenuation correction. The image reconstruction will start immediately after the 3D continuous acquisition, and if a motion artifact is detected, another 30 minutes continuous scan will be acquired.

### Structural Image Reconstructions

We used a newer version of the Freesurfer (Version 7) to re-process all the T1-weighted structural scans. Freesurfer [24] is an automated segmentation and cortical parcellation software package to reconstruct T1-weighted structural scans [25, 26]. All participants’ cortical surfaces are visually inspected/corrected by a demographic-blind technician. In the case of discrepancy, manual editing of the white and gray matter borders was conducted per the FreeSurfer manual editing guidelines [27]. Freesurfer segments the cortex into 33 different gyri/sulci-based regions in each hemisphere according to the Desikan-Killiany atlas [28],, and calculates the cortical thickness at each vertex, which is at the millimeter-by-millimeter resolution. The maps produced can detect submillimeter differences between groups. In addition, the sub-cortical regions are also segmented using a Bayesian classification technique to give 37 subcortical regional masks. The vertex-wise data are not constrained to the pre-defined ROIs and can be transferred to standard space using surface-based non-linear registration. To detect effects that are smaller in size or span over two or more pre-defined ROIs, utilization of the vertex-wise data is essential. The Freesurfer cortical and subcortical regions, as well as vertex-wise surface reconstruction, will be used to perform native space analysis of the fMRI and PET data. Finally, all the vertex-wise quantification of the Aβ, Tau and their overlap, as well as the surface-based probabilistic atlases will be done using Freesurfer reconstructed surfaces.

### Quantification Process for PET Data

An in-house develop fully automatic quantification method has already been implemented and evaluated using histopathological data and used in numerous published studies for the quantification of PET scans. Briefly, it starts by aligning dynamic PET frames (6 frames in Tau-PET and 4 frames in Amyloid-PET) to the first frame using rigid-body registration and averaging them to generate a static PET image. The PET image is then registered with the CT and merged to obtain a composite image in the PET static space. Each individual’s structural T1 image in FreeSurfer space is also registered to the same participant’s CT/PET composite image using normalized mutual information and 6 degrees of freedom to obtain a rigid-body transformation matrix to transfer all FreeSurfer regional masks to static PET image space. These regional masks in static PET space are used to extract the regional PET data. The standardized uptake value (SUV), defined as the decay-corrected brain radioactivity concentration normalized for injected dose and body weight, is calculated at selected regions, and then normalized to cerebellum gray matter to derive the standardized uptake value ratio (SUVR). SUVR will be determined at both the voxel and ROI levels.

The substantial and strong spill in a signal from white matter’s non-specific binding in Florbetapir is addressed by discarding the uptake in the gray-matter voxels located adjacent to white-matter volume both in the cerebellum for computing the reference region uptake and in the cerebral cortex for obtaining cortical regions’ SUVR. For AV1451 non-specific bindings are mainly in meninges, thus the same approach is used to discard all voxels adjacent to the surface of the gray-matter. For the surface reconstruction of the PET data, we sampled the Florbetapir on the gray-matter surface and AV1451 on the white-matter surface to address the spill-in issue from non-specific bindings. While these remedies helped compute the regional SUVRs the vertex-wise SUVRs, which are needed for obtaining the topographical overlapping of the two pathologies, were still significantly demonstrating a strong spill in signal. To completely address the issue, we developed a partial volume correction method that took advantage of the available scans from young and healthy participants and removed the spill-in signal completely from the PET scans.

### Partial Volume Correction

We developed simple but effective anatomy-driven partial volume correction. Each gray matter voxel’s uptake is a combination of actual binding in that location and the spill in from white matter non-specific binding. Using Florbetapir scans from 8 young (<30 years) and healthy participants we were was able to estimate the white matter spill in the signal. Since extremely young and healthy participants are not supposed to have any amyloid accumulation, any gray matter uptake in these subjects can be considered as a pure result of the spill in from adjacent regions with non-specific binding. In each young subject, we used the white-matter mask to extract the spatial distribution of the non-specific binding within the white matter region. Convolving this mask with the scanner point-spread function simulates the effect of the spill in signal perfectly since within the gray matter regions the correlation between actual uptake and the synthesized spill-in was more than 80%. Then we fitted a linear regression model for each voxel to predict the gray-matter spill using the synthesized spill in. Then using the fitted model parameters at each voxel, we estimated the gray matter spill in given the synthesized spill in and subtracted it from the actual gray matter uptake to completely remove it artifacts. So, without PVC all the sulci show a very high uptake due to white-matter nonspecific binding whereas it was completely removed in the corrected data.

### Topographical Overlapping of Aβ and Tau

To obtain topographical overlapping of the Aβ and Tau as well as generating a probabilistic atlas for the Tau, and Aβ uptake as well as their overlap it is required to transfer that PET data to standard space (MNI152). For vertex-wise analysis, we project the surface bases reconstructed Aβ and Tau to the surface of the MNI152 using spherical surface registration in FreeSurfer. Then setting a threshold (SUVR>1.3), we delineated vertices that have significant uptake of Aβ, Tau, or both pathologies. The subject-wise binary masks of the delineated vertices not only give the extend and expression (level) of each pathology as well as their overlap, but it can also be used to generate the probabilistic atlas for each pathology and their overlap. To quantify the extent and expression of each pathology within DMN, we combined 10 bilateral Freesurfer anatomical masks/labels (middle temporal, inferior parietal, lateral orbito-frontal, superior frontal, medial orbito-frontal, rostral middle frontal, precuneus, posterior cingulate, isthmus cingulate, and hippocampus) to generate a single binary mask to distinguish the vertices fall within DMN.

### Statistical Analysis

All statistical analyses and their visualization in this study are performed using Python and its main numeric modules numpy and matplotlib. ROC curves are computed using in-house developed python scripts and all the student t-tests and ANOVA are performed using SciPy statistical package, v6.1.1. The Chi-square test is performed using MATLAB v9.9.

## Results

Table I lists the number and demographic of the subjects used in this study for all three groups (HC, MCI, AD). Our subject selection was only based on the availability of the imaging data in the ADNI dataset within less than one year between data acquisitions in each modality and successful processing and analyzing the imaging data.

**Table 1:** Demographics of the subject used in the study

|  | HC-NC<sup>a</sup> | HC-C<sup>b</sup> | MCI-NC<sup>c</sup> | MCI-C<sup>d</sup> | AD<sup>e</sup> | P-value |
| --- | --- | --- | --- | --- | --- | --- |
| Number | 147 | 12 | 113 | 14 | 17 |  |
| Male/Female | 60/87 | 5/7 | 68/45 | 6/8 | 12/5 | 0.01<sup>f</sup> |
| Age (Mean ± STD) | 74.56 ± 7.58 | 83.33 ± 4.52 | 74.64 ± 7.16 | 75.32 ± 8.71 | 74.92 ± 9.36 | 0.004<sup>g</sup> |

<sup>a</sup> Healthy Controls – Non-Converters/ <sup>b</sup> Healthy Controls – Converters/ <sup>c</sup> Mild Cognitive Impairment – Non-Converter/ <sup>d</sup> Mild Cognitive Impairment – Converter/ <sup>e</sup> Alzheimer's Disease/ <sup>f</sup> Chi-square test/ <sup>g</sup> AVOVA test

ANOVA test revealed that there is a significant difference in the age of the 5 groups listed in Table one, the post hoc t-test determined that only the age of the converters from HC to MCI are significantly higher than the rest of the group. There were significantly more male participants in the MCI and AD group whereas the number of females was higher in both converters as well as HC groups.

Using 17 AD patients’ data in our dataset, we first show that the probability of observing overlapping Tau and Aβ pathologies within the DMN is significantly greater than observing it outside DMN, hinting at their more influential role in the pathophysiology of the AD. If the overlapping Aβ and Tau pathologies within the DMN are the true underlying pathophysiology of the AD, then all AD patients should have it. To show this, we first generated a vertex-wise probabilistic atlas for AD group where the value at each vertex gives the probability of observing overlapping Aβ and Tau in that group. Then, simply comparing the probabilities of the DMN vertices with the rest of the brain, will determine whether observing the overlapping pathologies within the DMN is more likely than outside DMN. Figure 1 shows the results of this examination. Figure 1a shows the probabilistic atlas of observing the overlapping pathologies obtained from AD group and overlaid over a semi-inflated cerebral cortex surface reconstructed from the MNI152 template. The probabilities are color-coded with hot colors where darker-red and red colors indicate lower probabilities and bright-red and yellow indicate higher probabilities. As it is seen, the probability of observing overlapping Aβ and Tau pathology is much higher in the DMN regions in comparison to the rest of the brain. Figure 1b quantifies this probabilistic atlas using boxplot and student t-test for determining the significant differences. Each boxplot illustrates the distribution of the probabilities for vertices that fall within DMN and compare it with the one that falls outside the DMN. Two hemispheres are plotted separately. Both right and left hemispheres show a significantly (right; t=132.73, p<0.0; left: t=112.20, p<0.0) higher probabilities of observing overlapping pathologies within the DMN in comparison to outside DMN. Therefore, preliminary evidence that overlapping pathology within DMN plays a more significant role in the underlying pathophysiology of the AD.

**Figure 1:**
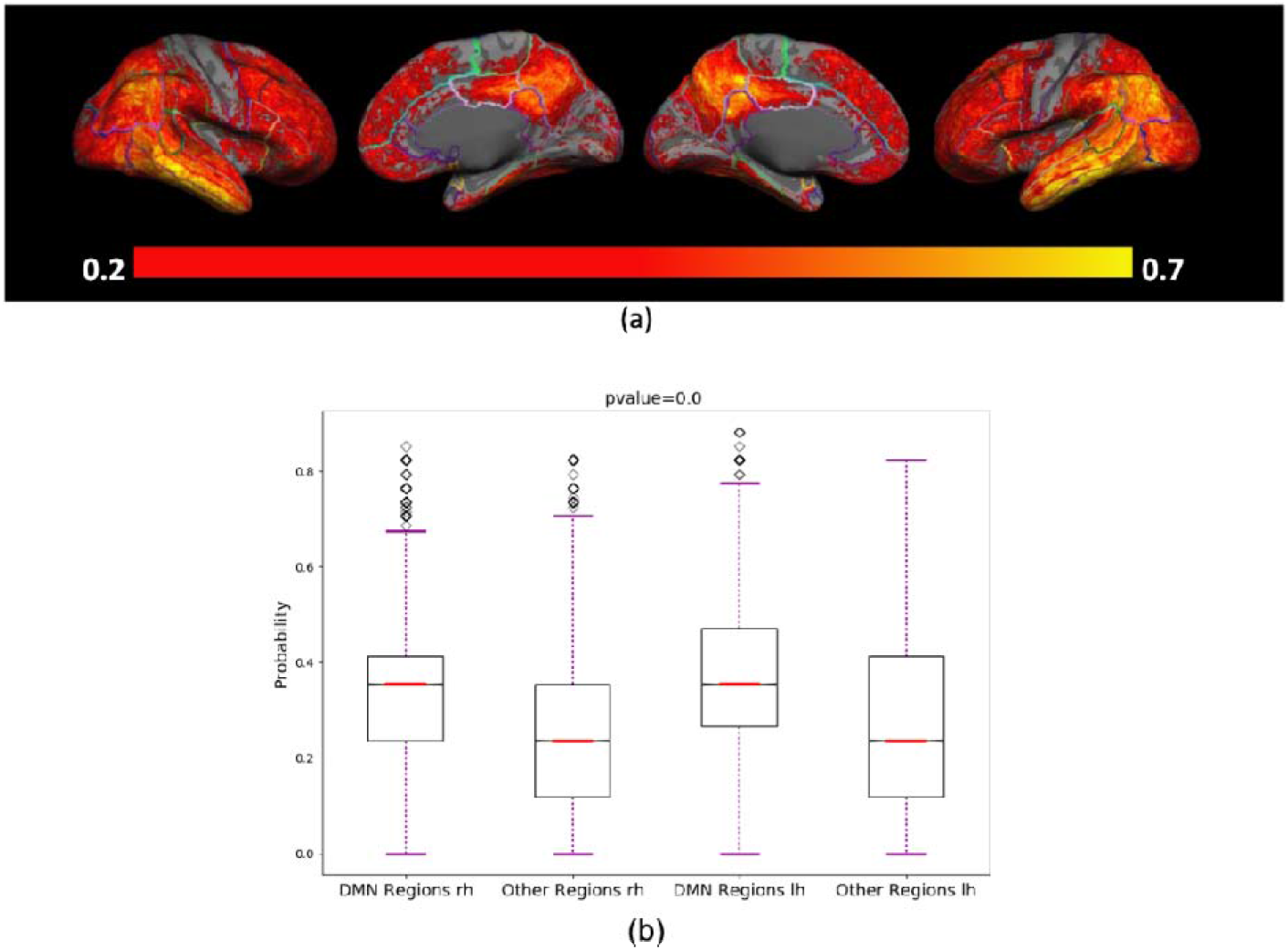
(a) The vertex-wise probabilistic atlas of observing overlapping pathologies throughout the entire cerebral cortex obtained from 17 AD patients. The probabilities at each vertex are color-coded with hot colors and overlaid over a semi-inflated cortical surface of the MNI152 template. (b) boxplots depict the distribution of the probabilities for vertices that fall within the DMN as well as the vertices that fall outside the DMN separately for each hemisphere.

Next, we show that in comparison to Aβ, Tau, and both pathologies, the overlapping pathology within the DMN has more predictability power to identify individuals converting from HC to MCI as well as from MCI to AD. We first start by creating a probabilistic atlas for Aβ and Tau pathology as well as their overlap for the HC and MCI groups. In addition, we generate a separate probabilistic atlas for individuals that are converting from HC to MCI and from MCI to AD. Figure 2 depicts these probabilistic atlases. Top row shows the probabilistic atlas for Aβ accumulation in the HC (left) and MCI (right) groups along with their converter which is depicted underneath. Middle row shows the same for Tau accumulation and bottom row illustrate the same for overlapping Aβ and Tau. It is clear, even visually, that overlapping Aβ, and Tau has more specificity for predicting the converters to MCI. While both Aβ and Tau have very high sensitivity (true positive) for detecting the converters, they have extremely poor specificity (true negative) to identify non-convertors. Note almost no overlapping pathology for HC non-converters and very minimal overlapping pathology in the MCI converters, suggesting the high specificity of the overlapping pathology to predict conversion from HC to MCI as well as from MCI to AD.

**Figure 2:**
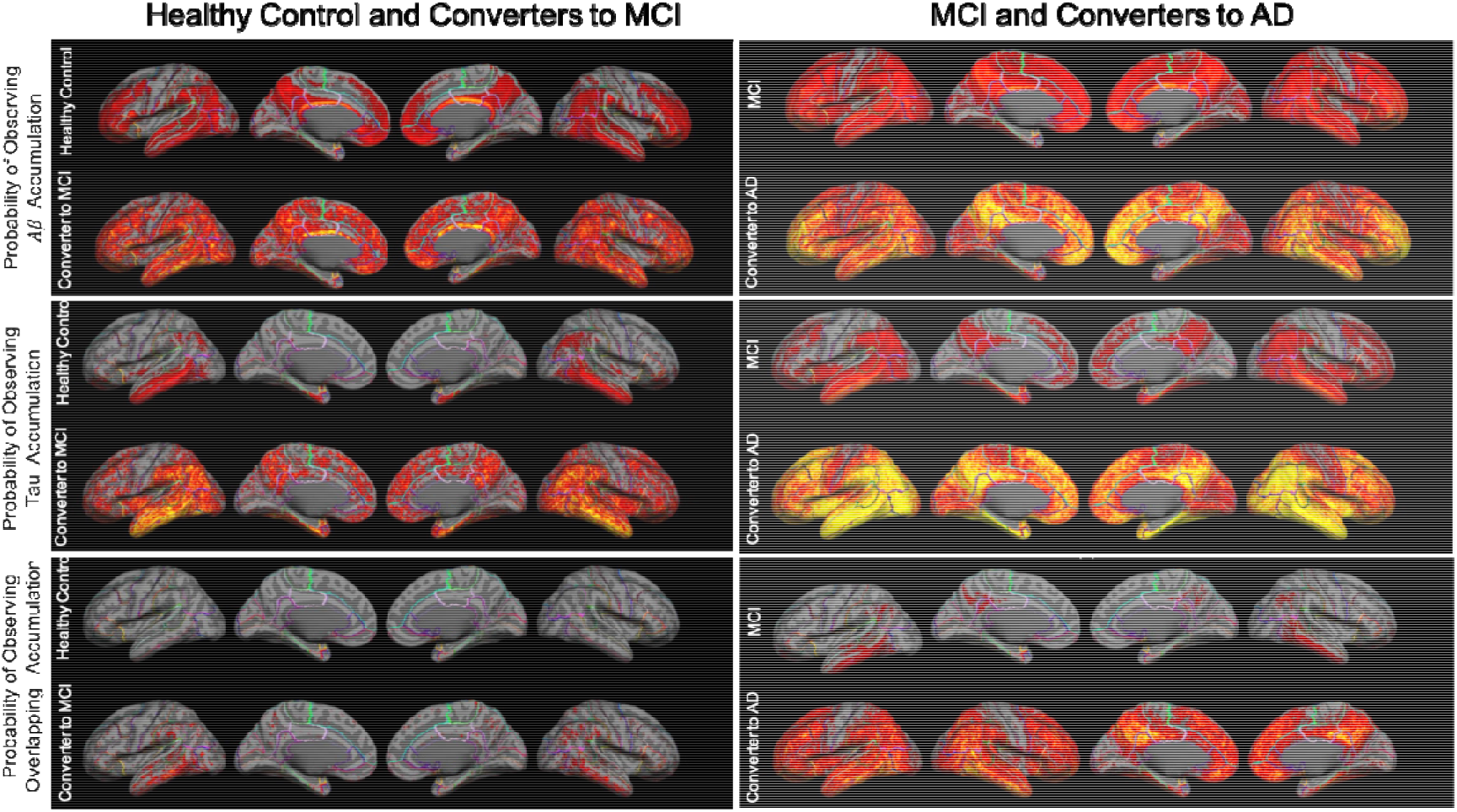
Vertex-wise probabilistic atlas for Ab (top row), Tau (middle row), and their overlap (bottom row) for 159 subjects in HC group (left) and 127 participants in MCI (right) group. The converters to MCI and AD are illustrated underneath each probabilistic atlas. The probabilities at each vertex are color-coded with hot colors and overlaid over a semi-inflated cortical surface of the MNI152 template.

To quantify the findings in Figure 2 we use ROC curves. Briefly, we set different thresholds for the extent of Aβ, Tau, and their overlap to detect the converters, and compute the true-positive and false-positive rate of each classification and plot the true-positive rates in terms of false-positive rates. Figure 3 shows the resultant ROC curves for (a) predicting the HC to MCI conversion using the whole brain and (c) considering only the DMN regions; (b) predicting MCI to AD conversion using the whole brain, and (d) using only DMN accumulation. As it is seen in Figure 3a, the predictability of the Aβ, Tau, and their overlap changes significantly across a different range of the false-positive rate. For higher false-positive rates the overlapping Aβ and Tau outperforms the predictability of both Aβ and Tau whereas for the lower false-positive rates its superiority reduces to a level lower than Tau predictability. We speculate that this loss of predictability in the lower range of false-positive rate is due to the massive accumulation of Aβ b and Tau in most of our participants. The same behavior can also be seen in the MCI to AD conversion as well as when we concentrate only to the DMN regions. The only difference is that for MCI participants the outperformance of the overlapping Aβ and Tau reduces significantly to the same level of Tau and Aβ. Again, we argue this is mainly because these subjects already passed the stage where we can accurately predict the conversion.

**Figure 3:**
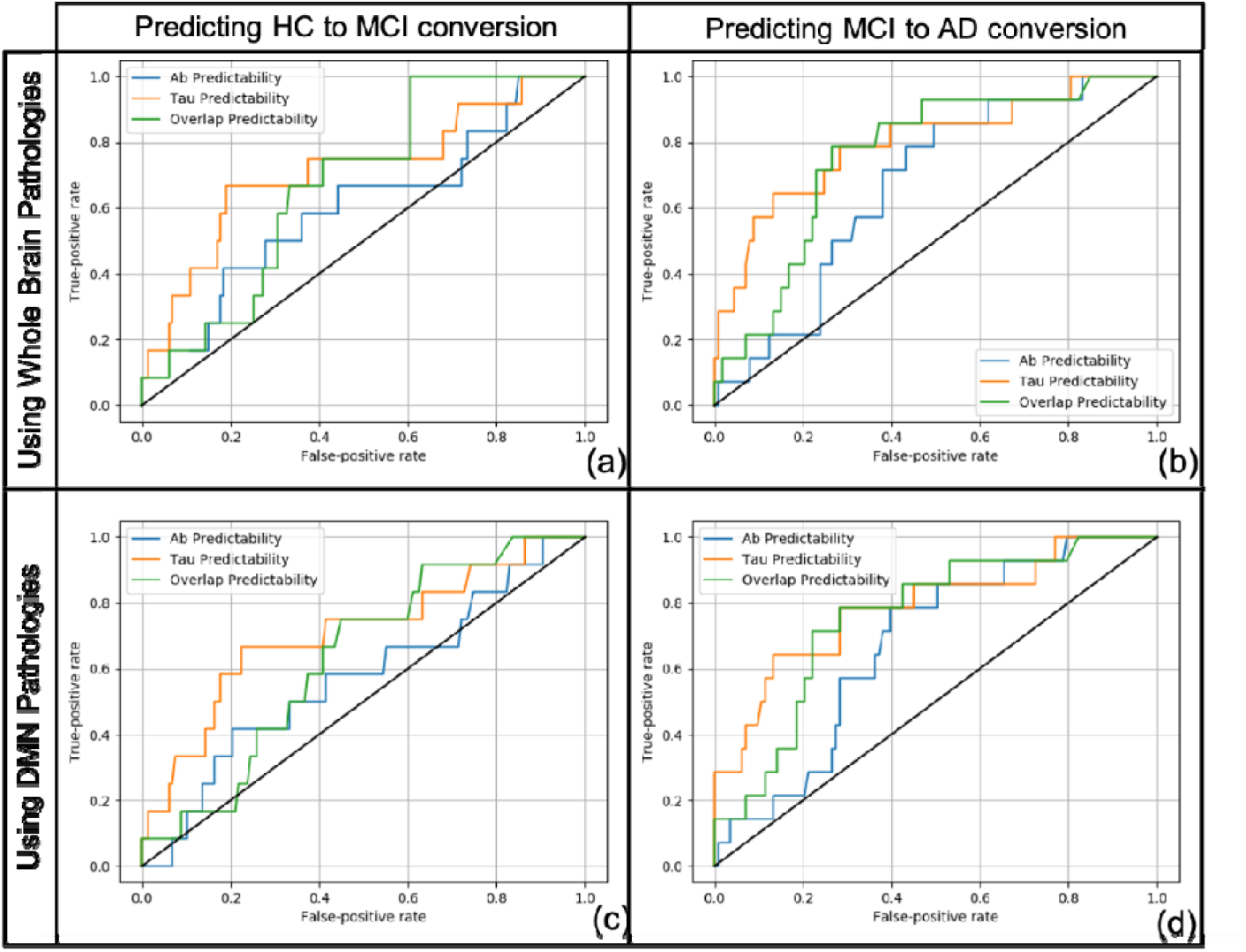
ROC curves obtained for predicting conversion (a&c) from HC to MCI and (b&d) from MCI to AD using accumulation of Ab, Tau, and their overlap (a&b) throughout the entire cerebral cortex and (c&d) when they are bounded to the DMN regions.

Finally, we demonstrate that the probability of observing overlapping pathology within DMN is much higher than the rest of the brain in HC and MCI converters, suggesting the more influential effect of overlapping Aβ and Tau when it occurs within DMN. Figure 4 shows the results of this analysis using boxplots and students t test. As seen, the probability of observing overlapping Aβ and Tau in the DMN is significantly higher when it is compared with the rest of the brain in both participants converting from (a) HC to MCI (t=79.90, p<0.0) and converters from (b) MCI to AD (t=227.14, p<0.0). This result suggests that the overlapping Aβ and Tau in the DMN is more influential for conversion from HC to MCI or from MCI to AD.

**Figure 4:**
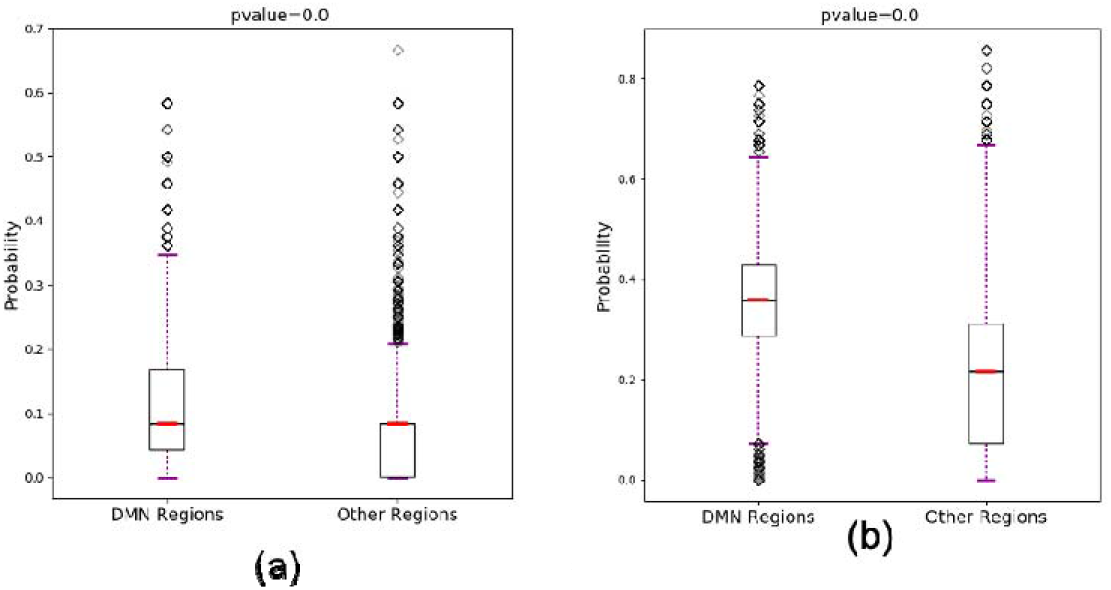
Boxplots illustrate the distribution of the vertex-wise probabilities of observing both Aβ and Tau for vertices within the DMN and in comparison with distribution of the probabilities of vertices falling outside DMN for participants (a) converting from HC to MCI and (b) from MCI to HC.

## Discussion

We have introduced the *double-insult* hypothesis in this work, where instead of Aβ and Tau pathology alone, the interaction of these two pathologies within the DMN plays the critical role in the underlying pathophysiology of AD. Using publicly available dataset (ADNI) and available *in-vivo* imaging data of the two pathologies from 303 participants (17 AD, 127 MCI, and 159 HC) which were quantified using our newly developed and evaluated technique, we provided preliminary evidence in support of our hypothesis. We first showed that within 17 AD patients, the probability of observing overlapping Aβ and Tau pathologies is significantly higher within DMN in comparison to the rest of the brain. Next, we showed that at least in the high false-positive range the true positive rate of detecting conversion from HC to MCI as well as MCI to AD has significantly increased. We argued that the reason such superiority is diminishing in the lower range of the false positive rate is because of massive Aβ and Tau uptake in some of the non-converter participants suggesting that there might be more converters in the HC group that has yet to be clinically diagnosed with AD. Future work is required to test our hypothesis on a younger population with a lower level of Aβ and Tau accumulation.

Alzheimer’s disease is a progressive neurodegenerative disease that is characterized by the presence of Aβ plaques and Tau tangles. Since these two pathologies can be initiated independently at different times and regions of the brain, how the accumulation of these two individual proteinopathies together causes enough neurological damage to develop AD is an unresolved challenge[14]. Recent reports suggest that at some point the independently initiated Tau and Aβ start interacting with each other, which subsequently accelerates not only the deterioration in their associated brain measurements but also the spread of the pathologies to other regions[14]. Thus, we hypothesis that it is the effects of this coupling between the two proteins that could potentially initiate a cascade of events that eventually results in significant neurodegeneration and cognitive decline. To understand the coupling effect, in the current study for the first time, we assess whether the interaction and overlap between Aβ and Tau tangles significantly accelerate the accumulation of both pathologies to starts the appearance of the disease symptoms. We examined the conversion probability of a relatively healthy participant to MCI or even MCI participant to AD when one proteinopathy or both are individually present in the DMN and compared it with the probability of observing the same pathology outside the DMN. In this study, we aimed at exploring the spatial distribution and overlap of hyperphosphorylated tau aggregation with fibrillar Aβ, to determine and effectively predict the conversion of the examined cognitively healthy subjects to MCI and MCI subjects to AD. We have shown that individuals with topographically overlapping Aβ and Tau accumulation within the DMN regions are more likely to convert to MCI or AD than the subject that has equivalent levels of but non-overlapping pathologies.

Both MCI and HC participants who converted to AD had a higher overlap inside their DMN regions. Therefore, the presence of overlap between Tau tangles and <u>Aβ</u> depositions inside the DMN region of the MCI and cognitively normal subjects can predict conversion to AD, whereas the presence of Tau or Amyloid only in the DMN region is not a powerful predictive factor of conversion. We used ROC curves to find the predictive factor of conversion by comparing the converters and non-converters. What this experiment demonstrated was that the extent of the p-Tau and Aβ pathologies in the brain, even if equal, does not signify that the person will convert to AD. The key component to the conversion lies in not only their overlap but in the location of the overlap.

One approach for disentangling spatiotemporal discrepancies is to trace the spatial and temporal characteristics of the documented AD pathologies from very early adulthood and study their relationship with other brain biomarkers and cognitive and behavioral measurements, which is only made possible by the recent advancement in the in-vivo imaging of brain functionality and AD pathologies[18, 29]. This approach has been adopted by ADNI, however, the lack of precise localization and quantification methods prevented them to take full advantage of these revolutionary in-vivo imaging techniques. In addition, sampling a population at the later adult life span essentially precludes them to capture the full dynamic of the AD pathology and its relationship with other brain biomarkers and cognitive and behavioral measurements. More specifically, the full dynamics of the brain large-scale networks and their spatiotemporal relationship with the two pathologies provides unprecedented insight about brain normal functioning as well as malfunctioning on a millimeter-by-millimeter scale and throughout the entire human cerebral cortex. Only through such precise quantification, we were able to investigate the effect of Aβ and Tau on the functionality of the Default mode network. Our recent findings show that DMN has two dissociable functional levels that are topographically overlapping but have distinct roles in the functional architecture of this large-scale brain network[30]. These findings coupled with the experiment outlined in this study indicates that the attenuation in the DMN’s functional connectivity (FC) and negative BOLD response (NBR) is significantly stronger for the overlapping accumulation of Aβ and Tau within DMN in comparison to the same level of non-overlapping Aβ and Tau pathologies, suggesting that double-insult to the functional structure of the DMN causes a complete breakdown of the network.

Our data on the topographic analysis of Tau tangles and Aβ fibrillation also presents further evidence that tauopathy can occur independently, as 24 subjects in our database had only p-Tau accumulation, without an underlying presence of AB pathology. This finding contrasts with previous studies that suggested that the development of a Tau tangle is contingent upon the presence of Aβ pathology and is generated dependently.

The pathophysiology of AD is still unclear, and there is an essential need to establish methods for predicting the progression of normal and MCI subjects to AD. This method will be clinically useful in detecting high-risk populations when they have the neuropathological changes in their brain and before their clinical symptoms of AD appear. Early detection of AD brain pathologies is of particular concern when new treatment methods have higher efficacy on early detected cases of AD.

## Conclusion

In conclusion, the present study demonstrates that the topographically overlapping pathologies, and not the existence of each AD proteinopathy alone, is a more accurate biomarker of AD, thus it is a more reliable predictor of a cognitively normal or MCI subject converting to an AD patient when the overlap is within the default mode network region of the brain. These findings shed light on the underlying pathophysiology of AD and pave the way to introduce a more accurate tool for not only the early detection of AD but also for a diagnosis path that can be investigated in future studies. Furthermore, this finding is valuable in developing disease-modifying treatments to slow down AD’s progression into an asymptomatic and fully developed disease.

## Ethical approval

“All procedures performed in studies involving human participants were in accordance with the ethical standards of the institutional and/ or national research committee and with the 1964 Helsinki declaration and its later amendments or comparable ethical standards.”

## Informed consent, data access and confidentiality of the data

According to ADNI protocol, “Informed consent was obtained in accordance with US 21 CFR 50.25, the TriCouncil Policy Statement: Ethical Conduct of Research Involving Humans and the Health Canada and ICH Good Clinical Practice.”

“Patient confidentiality was strictly held in trust by the participating investigators and research staff. This confidentiality was extended to cover testing of biological samples and genetic tests in addition to the clinical information relating to participants. All data will be transmitted securely via the Internet to ATRI at USC. Access is granted to study team members based on role.” “Data transmission will occur through a secure internet connection-https (hypertext transfer protocol secured) at 128-bit SSL. The study protocol, documentation, data and all other information generated will be held in strict confidence. No information concerning the study or the data will be released to any unauthorized third party, without prior written approval of the sponsoring institution.”

“All ADNI data are shared without embargo through the <u>LONI Image and Data Archive</u> (IDA), a secure research data repository. Interested scientists may obtain access to ADNI imaging, clinical, genomic, and biomarker data for the purposes of scientific investigation, teaching, or planning clinical research studies. Access is contingent on adherence to the ADNI Data Use Agreement and the publications’ policies outlined in the documents listed below. *Note: documents are subject to updates by ADNI*. The application process includes acceptance of the <u>Data Use Agreement</u> and submission of an online application form. The application must include the investigator’s institutional affiliation and the proposed uses of the ADNI data. ADNI data may not be used for commercial products or redistributed in any way.”

## Supporting information

data sharing approval from ADNI

## Data Availability

Data used in this paper were obtained from the Alzheimer's Disease Neuroimaging Initiative (ADNI) database (http://ADNI.loni.usc.edu). The investigators within the ADNI, who can be found at http://ADNI.loni.usc.edu/study-design/ongoing-investigations, contributed to the design and implementation of ADNI and/or provided data but did not participate in analysis or the writing of this article.

## Acknowledgments

Data used in this paper were obtained from the Alzheimer’s Disease Neuroimaging Initiative (ADNI) database (http://ADNI.loni.usc.edu). The investigators within the ADNI, who can be found athttp://ADNI.loni.usc.edu/study-design/ongoing-investigations, contributed to the design and implementation of ADNI and/or provided data but did not participate in analysis or the writing of this article. Data collection and sharing for this project were funded by the Alzheimer’s Disease Neuroimaging Initiative (National Institutes of Health Grant U01AG024904). ADNI is funded by the National Institute on Aging, the National Institute of Biomedical Imaging and Bioengineering, and through generous contributions from the following: AbbVie, Alzheimer’s Association; Alzheimer’s Drug Discovery Foundation; Araclon Biotech; Bio Clinical, Inc.; Biogen; Bristol-Myers Squibb Company; CereSpir, Inc.; Eisai Inc.; Elan Pharmaceuticals, Inc.; Eli Lilly and Company; EuroImmun; F. Hoffmann-La Roche Ltd and its affiliated company Genentech, Inc.; Fujirebio; GE Healthcare; IXICO Ltd.; Janssen Alzheimer Immunotherapy Research & Development, LLC.; Johnson &Johnson Pharmaceutical Research & Development LLC.; Lumosity; Lundbeck; Merck & Co., Inc.; Meso Scale Diagnostics, LLC.; NeuroRx Research; Neuro track Technologies; Novartis Pharmaceuticals Corporation; Pfizer Inc.; Piramal Imaging; Servier; Takeda Pharmaceutical Company; and Transition Therapeutics. The Canadian Institutes of Health Research is providing funds to support ADNI clinical sites in Canada. Private sector contributions are facilitated by the Foundation for the National Institutes of Health (www.fnih.org). The grantee organization is the Northern California Institute for Research and Education, and the study is coordinated by the Alzheimer’s Disease Cooperative Study at the University of California, San Diego. ADNI data are disseminated by the Laboratory for Neuro Imaging at the University of Southern California.

