## Supplementary material for "Topographical overlapping of the Aβ and Tau pathologies in the Default mode networks predicts Alzheimer’s Disease with higher specificity": data sharing approval from ADNI

### [Ext] Database Access Request

Fri 5/29/2020 10:10 AM

Congratulations. Your request for access to the Alzheimer's Disease Neuroimaging Initiative (ADNI) Data has been approved. If you already had a LONI user account your permissions have been updated to provide you access to ADNI data. If you did not yet have an account, an account will be created for you and an e-mail with your account information will be sent to you shortly.

Login page: https://ida.loni.usc.edu/login.jsp?project=ADNI&page=HOME
